# Mental health literacy and its correlates among Zimbabwean health sciences students: a cross-sectional study and exploratory psychometric evaluation

**DOI:** 10.64898/2026.09.16.26363279

**Authors:** Jacqueline Chindondondo, Odette Maungwa, Cathrine Chimutsa, Wendy Makoni, Mazvita W. Tapfumanei, Alfred Hove, Tariro D. Tunduwani, Sidney Muchemwa, Beatrice K. Shava, Shalom R. Doyce, Anotida R. Hove, Sandra Mboweni, Webster Mavhu, Dixon Chibanda, Jermaine M. Dambi

## Abstract

Globally, young adults, particularly university students, experience a high burden of mental disorders and multiple barriers to care, including limited mental health literacy (MHL). This study aimed to quantify MHL and associated factors among Zimbabwean health sciences students and, secondarily, to conduct an exploratory psychometric evaluation of the Mental Health Literacy Questionnaire (MHLq). This cross-sectional survey recruited 601 health sciences students using multistage sampling. MHL and common mental disorder (CMD) symptoms were measured using the 29-item MHLq and Shona Symptoms Questionnaire (SSQ-14), respectively. The MHLq total score was analysed continuously using multivariable ANCOVA with heteroscedasticity-robust standard errors. Exploratory psychometric analyses assessed internal consistency, factorability, factor structure and hypothesis-based construct validity. The mean MHLq total score was 119.5 (SD = 10.6). The total scale showed good internal consistency (Cronbach‘s α = 0.85; McDonald‘s ω = 0.87), although domain-level reliability was lower (α = 0.52–0.66). Sampling adequacy for factor analysis was good (KMO = 0.87; Bartlett‘s test p < 0.001); parallel analysis suggested five empirical factors, and the original four-domain structure was only partially reproduced. Construct-validity analyses showed higher MHLq scores among students exposed to psychology/psychiatry courses, students in later years of study, and those with higher digital literacy, while MHLq scores were only weakly related to CMD symptoms. In adjusted ANCOVA, psychology/psychiatry course exposure, digital literacy, study year and family/friend information sources were independently associated with MHLq score; drug/substance use and personal CMD history were associated with lower scores. The MHLq total score demonstrated good internal consistency and partial support for construct validity in this setting, but the weaker domain reliabilities and incomplete replication of the original factor structure support cautious interpretation of subscale scores. Analysing the total MHLq continuously is therefore preferable for the present study. Campus-wide mental health education and digital-literacy strengthening merit further evaluation in longitudinal and intervention studies.

## Introduction

Globally, 1 in every 8 people lives with a mental disorder [1]. The burden of mental disorders is disproportionately higher in low- and middle-income countries (LMICs) than in high-income countries; LMICs account for 80% of mental disorders globally [2]. However, mental illness awareness and treatment rates are comparatively lower in LMICs than in high-income countries [3]. For example, only 15–24% of persons requiring treatment for severe mental illness in LMICs have access to care [4]. The low treatment coverage and awareness are attributable to stigma, low mental health literacy, low prioritisation of mental services, and a lack of resources (e.g., a lack of qualified mental healthcare practitioners) [5].

Most common mental disorders (CMDs) emerge during early adulthood and, if untreated, can spill into adulthood, with multiple ripple effects [6]. For instance, 50% of mental health disorders in Africa develop by 14 years and 75% by 24 years [7]. Further, systematic reviews show a high global prevalence of CMDs in young adults, especially college/university students, ranging from 25% to 31% [8–10]. Unfortunately, young adults experience several barriers to mental health care, including low mental health literacy (MHL), stigma, and poor access to mental health facilities [11]. Of the existing barriers to mental healthcare, MHL is a salient predictor of mental health-seeking behaviours in young adults and is a potentially modifiable target for intervention [3,12].

Mental health literacy (MHL) has been defined as “knowledge and beliefs about mental disorders which aid their recognition, management or prevention” [13]. MHL is multidimensional and varies across cultural and contextual settings. For instance, previous studies have demonstrated variable MHL prevalence among university students, ranging from as low as 12% among Nigerian students to as high as 74% among Australian students [14,15]. Further, a recent systematic review demonstrated gender, year of study, faculty and ethnicity as salient predictors of MHL among university students [16]. For instance, females were twice as likely as males to have higher MHL (AOR = 1.72, 95% CI 1.26–2.34) [17]. It is hypothesised that females are likely to have stronger social support systems, specifically informational social support, that drives increased CMD awareness compared to males, and consequently higher MHL [2]. Secondly, programme of study has been associated with MHL, but this is not without controversy [17–20]. For example, a cross-sectional study (N = 4671) assessing depression literacy among undergraduates in Sri Lanka found that medical undergraduates were better at recognising mental health conditions and showed greater alignment with expert opinion on treatment options than students in other faculties, indicating high MHL [18]. Elsewhere, an American cross-sectional study of university students (N = 1213) found that applied health sciences students had MHL levels comparable to those of business/accounting students (15.3% vs. 14.0%) [17]. The equivalent MHL levels are noteworthy, as applied health sciences students are hypothesised to have higher MHL levels due to increased exposure to mental health-related knowledge from their training curricula [17]. Consequently, there have been calls to investigate and improve the MHL of healthcare students for several reasons [21]. Of note, healthcare students experience higher stress levels and poorer mental health functioning than non-healthcare students [22,23]. The reasons behind the high burden of CMDs among university students are multifaceted, ranging from significantly higher academic load, stigma, and financial constraints to poor MHL [22,24,25]. Indeed, the debilitating impacts of untreated CMDs on healthcare students are far-reaching and may have negative ramifications for their roles as future healthcare professionals. For instance, low MHL levels may lead to the propagation of stigmatising attitudes among healthcare professionals [26]. Early evidence-based interventions bolstered by high-quality epidemiological data are thus essential. The primary objective was to determine mental health literacy among Zimbabwean health sciences students and to identify associated factors to inform context-appropriate interventions. A secondary objective was to conduct an exploratory psychometric evaluation of the MHLq in this sample, focusing on internal consistency, structural validity and hypothesis-based construct validity. The study also provides baseline normative data against which future MHL-enhancement interventions can be assessed.

## Materials and methods

### Study design

This cross-sectional study was conducted from February to August 2023 in Harare, Zimbabwe. Recruitment ran from 27 February to 31 March 2023.

### Study setting

Participants were drawn from the University of Zimbabwe, Faculty of Medicine and Health Sciences, a standalone faculty at the Parirenyatwa Group of Hospitals in Harare, away from the University of Zimbabwe main campus. The Faculty of Medicine and Health Sciences houses over 2,000 students across 18 programmes, including Radiography, Medicine, Physiotherapy and Occupational Therapy.

### Study participants

Eligible participants were registered undergraduate students aged ≥18 years who were enrolled in a Faculty of Medicine and Health Sciences programme and had no previous formal psychiatric diagnosis. Students unable to provide informed consent were excluded.

### Sampling and sample size calculation

Using a 5% margin of error, 80% power, and an expected mean MHL score of 69.6 (SD = 7.8) based on a comparable Iranian university study [12] and a 10% allowance for non-response or incomplete data, a minimum sample of 597 participants was required. The sample size was calculated using the STATISTICA power function (version 14). Multistage sampling was used: participants were first stratified by degree programme and then recruited proportionately within strata using convenience sampling.

### Data collection

Participants were approached by JC, OM, CC, WM, MT, SRD and AH in lecture theatres, and the study’s aims were briefly explained to them. Students who volunteered were given the consent form and informed of their right to withdraw from the study at any time. After confirming understanding, participants provided written informed consent and were given electronic tablets to complete the self-administered questionnaire.

### Instrumentation

#### Demographic questionnaire

This questionnaire asked about age, sex, marital status, programme, year of study, religion and current smoking and alcohol consumption status. The questionnaire also asked about the availability of internet access and digital literacy, perceived financial status as a socioeconomic status proxy, and exposure to mental health disorders through personal, friend or family diagnosis.

#### Mental Health Literacy Questionnaire - young adult version (MHLq)

The MHLq is a 29-item Likert-scale questionnaire with responses ranging from 1 (strongly disagree) to 5 (strongly agree) [27]. It assesses four conceptual domains: knowledge of mental health problems, erroneous beliefs/stereotypes, help-seeking and first-aid skills, and self-help strategies. Six negatively keyed erroneous-belief items are reverse-scored so that higher item, domain and total scores consistently indicate higher mental health literacy; the total score ranges from 29 to 145 [27]. The initial young-adult validation reported Cronbach‘s α = 0.84 for the total scale and a four-factor structure [27]. The MHLq has also been translated and preliminarily validated in Chichewa for use in Malawi [28]. For the present analyses, domain scores were reconstructed from the published item-domain assignments, and reverse-keyed items were retained in their directionally scored form for scale analyses.

#### Shona Symptom Questionnaire (SSQ-14)

Developed in Zimbabwe, the SSQ-14 is a self-administered tool used to detect depression and anxiety. It consists of 14 binary-coded (yes/no) questions that ask participants whether they have experienced the following symptoms in the past week. Scores range from 0–14, with a cut-point of ≥9 indicating risk of CMDs. The tool was validated in a local primary care population, with internal consistency (Cronbach‘s α = 0.74) and, at the ≥9 cut-point, sensitivity of 84% and specificity of 73% for depression and/or general anxiety disorders [29].

### Ethical considerations

Ethical approval was granted by the Joint University of Zimbabwe and Parirenyatwa Hospital Research Ethics Committee (Ref: JREC/72/2023). The study was conducted in accordance with the Declaration of Helsinki. All participants provided written informed consent before participation, and study information was handled confidentially.

### Data analysis

Descriptive statistics (frequencies, percentages, means and standard deviations) were computed for participant characteristics and study outcomes. Because the MHLq has no validated diagnostic or clinical cut-off, the 29-item MHLq total score (range 29–145, with higher scores indicating greater mental health literacy) was analysed as a continuous outcome. Candidate determinants were first examined using unadjusted general linear models (t-tests/ANOVA or simple linear regression, as appropriate). Variables with p ≤ 0.20 were entered into a multivariable ANCOVA/general linear model; backward elimination was then used at the factor level while age and sex were retained a priori. Age was mean-centred. Sparse categories were combined where appropriate (very low/low digital literacy; very poor/poor perceived health). Homogeneity of regression slopes was assessed using age-by-factor interactions; the age-by-drug/substance-use interaction was significant and therefore retained in the final model. Residual diagnostics indicated heteroscedasticity, so HC3 heteroscedasticity-robust standard errors, confidence intervals and Wald tests are reported. Multicollinearity was not material (all variance inflation factors <5).

Secondary exploratory psychometric analyses were conducted on the MHLq in this sample. Internal consistency was estimated using Cronbach‘s α and McDonald‘s ω for the total scale and the four published conceptual domains. Factorability was assessed using the Kaiser-Meyer-Olkin (KMO) statistic and Bartlett‘s test of sphericity. Exploratory factor analysis was conducted using principal-axis factoring with oblimin rotation because correlated MHL dimensions were expected. The number of factors was guided by parallel analysis using 1,000 random permutations; a forced four-factor solution was also examined as a sensitivity analysis because the original MHLq describes four domains [27]. Hypothesis-based construct validity analyses, specified for this secondary analysis based on prior MHL literature, examined whether MHLq scores were higher with psychology/psychiatry course exposure, later study year, greater digital literacy, and proximity to someone with a mental disorder, and whether MHLq scores showed only a weak relationship with current CMD symptoms. Cohen‘s d, η^2^ and Pearson‘s r were reported as appropriate. These psychometric analyses were exploratory, and no multiplicity adjustment was applied. All tests were two-sided at α = 0.05. Analyses were performed in Python 3.13.5 using statsmodels 0.14.6 and SciPy 1.17.0.

## Results

### Participant characteristics

A total of 601 participants were recruited into the study; their mean age was 22.7 (SD = 1.7) years. Among participants, 57.9% were female, 41.4% were fourth-year students, 34.9% were medical students, 74.4% had taken a psychology/psychiatry course, 91.8% reported a religion that acknowledges mental health, 92.2% had optimal-to-very-high internet access, and 96.7% had optimal-to-very-high digital literacy. Regarding lifestyle factors, 8.5% smoked, 33.4% drank alcohol, and 7.3% reported drug/substance use. Overall, 59.2% rated their health status as good or very good. Personal history of a mental health disorder was reported by 3.8%, 20.8% reported a friend with a mental health disorder, and 28.3% reported a family member with such a history. A blended physical-and-digital mode was the preferred mental health service modality for 52.4% of participants. Participant characteristics are summarised in **Table 1**.

**Table 1.**
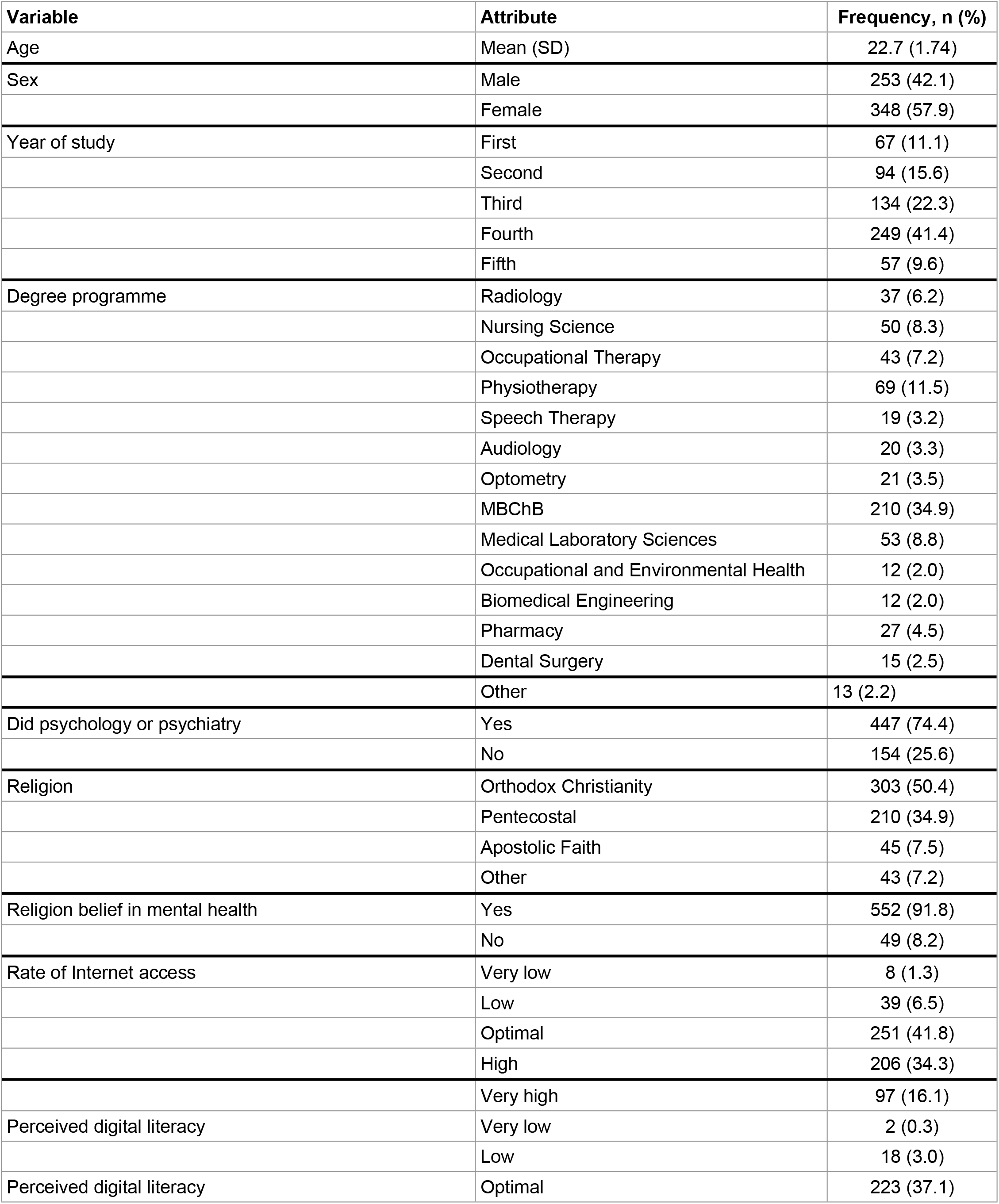
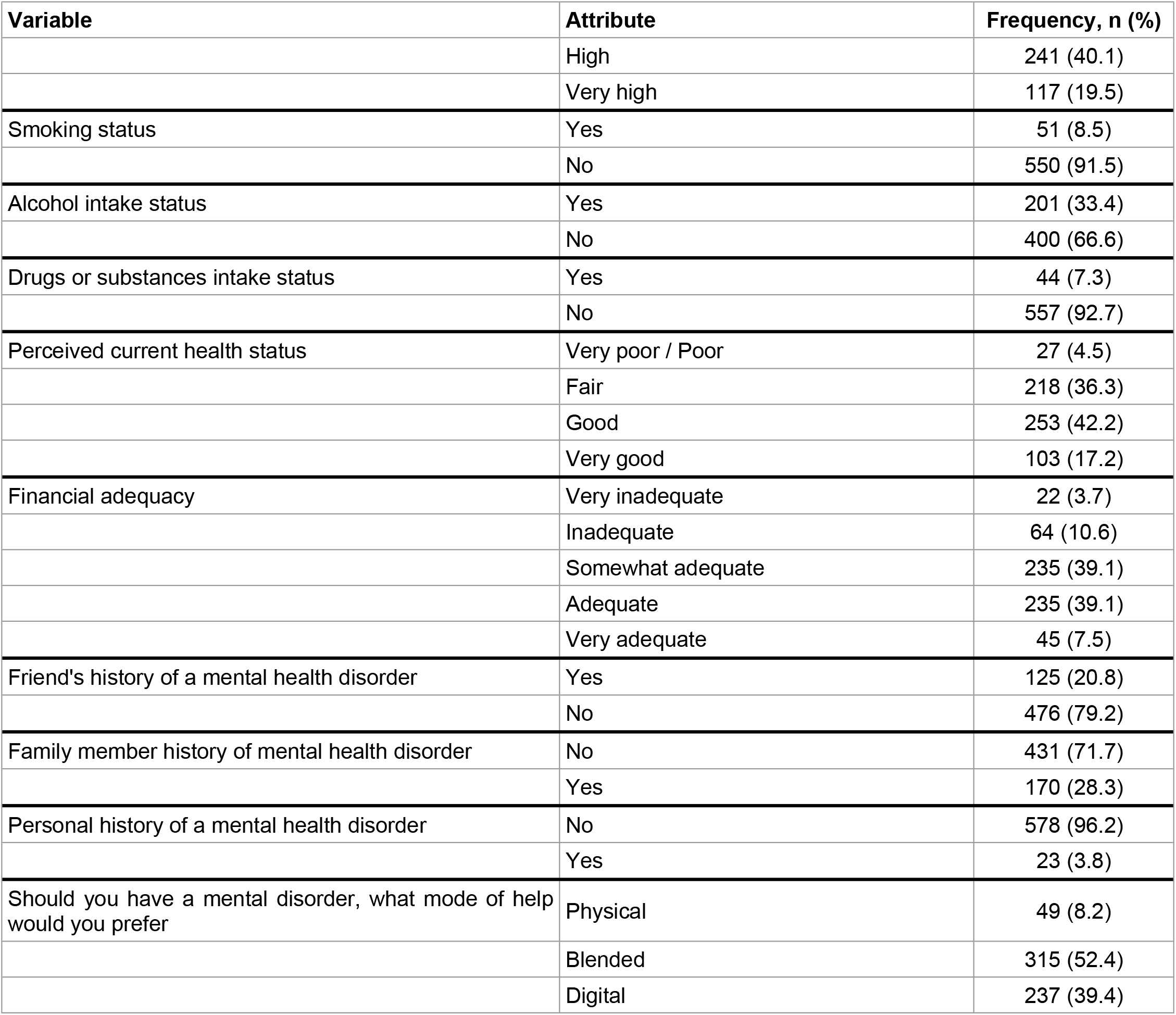
participant characteristics (n = 601)

### Information sources

The four most common sources of information were educational books (72.2%), healthcare staff (67.9%), WhatsApp (68.1%) and YouTube (64.6%) **(Table 2)**.

**Table 2.** Sources of mental health information (n = 601)

| Sources of information | Frequency, n (%) |  |
| --- | --- | --- |
|  | Yes | No |
| Healthcare staff | 408 (67.9) | 193 (32.1) |
| Magazines or newspapers | 202 (33.6) | 399 (66.4) |
| Educational books | 434 (72.2) | 167 (27.8) |
| Family or friends | 360 (59.9) | 241 (40.1) |
| Traditional healers | 28 (4.7) | 573 (95.3) |
| Television | 333 (55.4) | 268 (44.6) |
| Radio | 230 (38.3) | 371 (61.7) |
|  | Yes | No |
| Social media | 390 (64.9) | 211 (35.1) |
| Facebook | 358 (59.6) | 243 (40.4) |
| WhatsApp | 409 (68.1) | 192 (31.9) |
| Twitter | 238 (39.6) | 363 (60.4) |
| Instagram | 273 (45.4) | 328 (54.6) |
| TikTok | 267 (44.4) | 334 (55.6) |
| YouTube | 388 (64.6) | 213 (35.4) |
| WeChat | 19 (3.2) | 582 (96.8) |
| Tumblr | 7 (1.2) | 594 (98.8) |
| LinkedIn | 66 (11.0) | 535 (89.0) |
| Reddit | 11 (1.8) | 590 (98.2) |
| Other smartphone applications | 28 (4.7) | 573 (95.3) |

### Mental health literacy

Most participants (95.2%) agreed that highly stressful situations may cause mental health conditions, while 22.8% disagreed that people with schizophrenia usually have delusions. In addition, 94.3% disagreed that mental health disorders do not affect people‘s behaviours, and 72.9% disagreed that people with mental disorders belong to low-income families. Regarding help-seeking behaviour, 92.1% reported that they would seek help from a psychologist if they had a mental health condition, whereas 17.0% disagreed that they would encourage someone with a mental disorder to see a psychiatrist. In the self-help domain, 94.5% agreed that a balanced diet contributes to good mental health and 82.6% agreed that sleeping well contributes to good mental health **(S1 Table)**. Using the published conceptual item-domain assignments, the mean domain scores were knowledge of mental health problems, 45.0 (SD = 4.5); erroneous beliefs/stereotypes, 33.3 (SD = 3.4); help-seeking/first-aid skills, 24.5 (SD = 2.8); and self-help strategies, 16.8 (SD = 2.0). The mean total MHLq score was 119.5 (SD = 10.6; range 73–143) **(Table 3)**.

**Table 3.** Descriptive statistics for Mental Health Literacy Questionnaire scores (N = 601)

| Domain | Min. – Max. | Mean (SD) | Median [Q1 – Q3] |
| --- | --- | --- | --- |
| Knowledge of mental health problems | 30 – 55 | 45.0 (4.5) | 45 (42 – 49) |
| Erroneous beliefs/stereotypes | 18 – 40 | 33.3 (3.4) | 33 (31 – 36) |
| Help-seeking and first-aid skills | 14 – 30 | 24.5 (2.8) | 24 (23 – 26) |
| Self-help strategies | 6 – 20 | 16.8 (2.0) | 17 (16 – 18) |
| Total MHLq | 73 – 143 | 119.5 (10.6) | 119 (113 – 127) |

### Common mental disorders symptoms

The most reported psychiatric morbidity symptoms were overthinking (64.4%), lethargy (58.7%), and failing to concentrate (54.1%). Only 4.5% reported suicidal ideation, while 5.2% reported experiencing hallucinations **(S2 Table)**. Using an SSQ-14 cut-off score of ≥9, the prevalence of CMDs in this sample was 14.5%.

### Multivariable ANCOVA

Unadjusted associations between candidate determinants and the continuous MHLq total score are presented in **S3 Table**. The final multivariable ANCOVA explained 18.2% of the variance in MHLq scores (adjusted R^2^ = 0.158; robust F(17, 583) = 6.47; p < 0.001). Exposure to psychology/psychiatry courses was associated with a 3.59-point higher MHLq score (95% CI, 1.53 to 5.65; p < 0.001), and using family/friends as a mental health information source was associated with a 3.08-point higher MHLq score (95% CI 1.40 to 4.76; p < 0.001). Digital literacy was independently associated with MHLq score (overall p = 0.001): compared with very high digital literacy, low, optimal and high digital literacy were associated with scores lower by 9.39 (95% CI −16.46 to −2.32), 4.27 (95% CI −6.66 to −1.87) and 3.20 points (95% CI −5.44 to −0.97), respectively. Study year was also associated with MHLq score (overall p = 0.004); third-year and fifth-year students scored 3.53 (95% CI 0.31 to 6.75; p = 0.032) and 7.26 points (95% CI 2.66 to 11.86; p = 0.002) higher than first-year students, respectively. Perceived health status showed an overall association (p = 0.007), although the pattern was not monotonic: students reporting good health scored 4.03 points lower than those reporting very good health (95% CI −6.53 to −1.54; p = 0.002), whereas the poor and fair categories did not differ significantly. At the mean age of 22.7 years, drug/substance use was associated with a 6.78-point lower MHLq score (95% CI, −11.40 to −2.17; p = 0.004), with the size of this difference attenuating with increasing age (age × drug/substance-use interaction B = 2.72; 95% CI, 0.24 to 5.20; p = 0.032). A personal CMD history was associated with a 4.12-point lower score (95% CI, −8.22 to −0.02; p = 0.049). Sex and the centred age main effect were not independently associated with the MHLq score **(Table 4)**.

**Table 4.** Multivariable ANCOVA of factors associated with continuous MHLq total score (N = 601)

| Predictor | Comparison | Adjusted B (robust SE) | 95% CI | p-value | Overall p |
| --- | --- | --- | --- | --- | --- |
| Psychology/Psychiatry course | Yes vs No | 3.59 (1.05) | 1.53 to 5.65 | <0.001 | <0.001 |
| Digital literacy | Low vs Very high | -9.39 (3.60) | -16.46 to -2.32 | 0.009 | 0.001 |
|  | Optimal vs Very high | -4.27 (1.22) | -6.66 to -1.87 | <0.001 |  |
|  | High vs Very high | -3.20 (1.14) | -5.44 to -0.97 | 0.005 |  |
| Family/friends information source | Yes vs No | 3.08 (0.85) | 1.40 to 4.76 | <0.001 | <0.001 |
| Health status | Poor vs Very good | -1.23 (2.45) | -6.04 to 3.58 | 0.614 | 0.007 |
|  | Fair vs Very good | -2.16 (1.36) | -4.83 to 0.52 | 0.113 |  |
|  | Good vs Very good | -4.03 (1.27) | -6.53 to -1.54 | 0.002 |  |
| Drug/substance use | Yes vs No | -6.78 (2.35) | -11.40 to -2.17 | 0.004 | 0.004 |
| Study year | Second vs First | 1.48 (1.57) | -1.60 to 4.56 | 0.346 | 0.004 |
|  | Third vs First | 3.53 (1.64) | 0.31 to 6.75 | 0.032 |  |
|  | Fourth vs First | 1.99 (1.70) | -1.34 to 5.33 | 0.240 |  |
|  | Fifth vs First | 7.26 (2.34) | 2.66 to 11.86 | 0.002 |  |
| Sex | Male vs Female | -1.46 (0.89) | -3.21 to 0.30 | 0.104 | 0.104 |
| Personal CMD history | Yes vs No | -4.12 (2.09) | -8.22 to -0.02 | 0.049 | 0.049 |
| Age | Per 1-year increase | -0.45 (0.38) | -1.18 to 0.29 | 0.233 | 0.233 |
| Age × drug/substance use | Interaction | 2.72 (1.26) | 0.24 to 5.20 | 0.032 | 0.032 |

**Note:** B is the adjusted mean difference in MHLq points relative to the stated reference group. Age was mean-centred at 22.66 years; therefore, the drug/substance-use main effect is evaluated at the sample mean age. HC3 robust standard errors and Wald p-values are reported because heteroscedasticity was detected. Low digital literacy combines the very low/low categories; poor health combines the very poor/poor categories. Model $R^2 = 0.182$ ; adjusted $R^2 = 0.158$ ; robust $F(17, 583) = 6.47$ , $p < 0.001$ .

### Supplementary psychometric performance of the MHLq

The 29-item MHLq total score showed good internal consistency (Cronbach‘s α = 0.848; McDonald‘s ω = 0.866). Reliability was lower for the conceptual domains: knowledge α = 0.661, ω = 0.711, erroneous beliefs/stereotypes α = 0.626, ω = 0.654, help-seeking/first-aid α = 0.523, ω = 0.562, and self-help α = 0.515, ω = 0.535 (**S4 Table**). The item correlation matrix was suitable for factor analysis (KMO = 0.869; Bartlett‘s χ^2^ (406) = 4537.8, p < 0.001). Parallel analysis retained five factors: the first five observed eigenvalues (6.48, 2.33, 1.70, 1.50 and 1.32) exceeded the corresponding 95th-percentile random eigenvalues (1.48, 1.41, 1.36, 1.32 and 1.28), whereas the sixth did not. The oblique five-factor solution contained several cross-loadings and did not map cleanly onto the original four conceptual domains; a forced four-factor sensitivity solution likewise provided only partial structural replication **(S5 Table)**. These findings favour use of the overall MHLq score while cautioning against strong subscale-level interpretation in this sample. Hypothesis-based construct validity was mixed but generally supportive of the total score (**S6 Table)**. Students exposed to psychology/psychiatry courses had higher unadjusted MHLq scores than those without exposure (120.7 vs 116.0; Cohen‘s d = 0.46; p < 0.001). MHLq scores differed by year of study (F(4, 596) = 4.81, p < 0.001, η^2^ = 0.031), increasing from 117.0 in first-year students to 124.4 in fifth-year students, although the pattern was not strictly linear. Scores also differed across digital-literacy categories (F(4, 596) = 6.77, p < 0.001, η^2^ = 0.043). In contrast, having a friend or family member with a mental disorder was not associated with higher MHLq scores (p = 0.329 and p = 0.835, respectively). The association between MHLq and SSQ-14 symptom scores was weak (r = −0.073, p = 0.074), supporting discrimination between mental health literacy and current psychological distress.

## Discussion

### Synthesis

This study assessed mental health literacy among health sciences students, examined associated factors, and conducted secondary exploratory psychometric analyses of the MHLq. The mean MHLq total score was 119.5 (SD = 10.6). Exposure to psychology/psychiatry courses, higher digital literacy, study year, and using family/friends as an information source were associated with higher mental health literacy. Conversely, drug/substance use and personal CMD history were associated with lower scores; perceived health status was also associated with mental health literacy but without a simple dose-response pattern.

### Comparison of mental health literacy scores with past studies

Our mean MHLq score was higher than the 105.3 (SD = 7.1) reported in a validation study of Portuguese youth [30]. Differences in population, educational exposure and context may contribute to this variation. The supplementary psychometric findings strengthen the rationale for using the MHLq total score, while also identifying important measurement limitations. Total-scale internal consistency (α = 0.85) was essentially identical to the α = 0.84 reported in the original young-adult validation [27], and ω = 0.87 provided convergent evidence of reliability. However, the four domain scores showed only modest internal consistency (α = 0.52–0.66), lower than most coefficients reported in the original validation [27]. Parallel analysis also suggested a five-factor empirical solution, and neither the five-factor nor forced four-factor solution reproduced the published conceptual structure cleanly. Cross-cultural variation in item interpretation, educational exposure and contextual meanings of mental illness may contribute to this structural instability. Accordingly, the present findings should not be interpreted as a definitive local validation; rather, they support the total score for epidemiological comparison while indicating a need for dedicated Zimbabwean validation work, ideally including cognitive interviewing, confirmatory factor analysis in an independent sample and assessment of measurement invariance.

Previous research from LMIC and university settings has consistently shown substantial variation in mental health literacy, but direct comparisons are complicated by differences in instruments, scoring approaches and cut-offs. For example, an Ethiopian university study reported a 50.9% prevalence of higher MHL and identified digital health literacy, field of study and exposure to mental illness as relevant factors [31]. A Zimbabwean study among radiography students (N = 89), using a different Mental Health Literacy Scale, reported a mean score of 96.6 (SD = 9.6) [32]. Rather than assigning participants to an arbitrary high/low category, the present analysis provides a continuous normative estimate that may be more useful for monitoring change in future MHL-enhancement interventions.

### Factors associated with mental health literacy

#### Psychology or psychiatry course exposure

Students exposed to psychology or psychiatry courses had MHLq scores approximately 3.6 points higher than those without such exposure after adjustment. This is consistent with an American cross-sectional study of university students (N = 1213), in which exposure to psychology courses was strongly associated with mental health literacy [17], and with evidence from UK university students [33]. Educational curricula can improve knowledge of mental illness and reduce stigmatising attitudes [34,35]. These findings support consideration of introductory mental health content, including mental health first aid or equivalent structured modules, across healthcare programmes rather than restricting such training to disciplines with formal psychiatric content.

#### Year of study

Mental health literacy also varied by year of study. The largest adjusted difference was observed for fifth-year students, who scored 7.3 points higher than first-year students; third-year students also scored higher, whereas the differences for second- and fourth-year students were not statistically significant. This pattern supports an association between cumulative academic exposure and MHL, but it is not strictly linear. A systematic review likewise identified year of study as a correlate of MHL among university students [36]. Prior work in UK medical students similarly reported higher MHL scores in later years of training [37], potentially reflecting increasing exposure to mental health education and clinical learning [18,37,38]. The non-linear pattern in the present study suggests that specific curricular exposures, rather than year of study alone, should be examined in future longitudinal work.

#### Lived experience and interpersonal information sources

The continuous-score analysis refined the interpretation of lived experience and interpersonal information sources. Using family or friends as a source of mental health information was independently associated with a 3.1-point higher MHLq score, whereas simply having a friend or family member with a CMD history was not retained in the final model. In contrast to some published evidence [31,39,40], participants reporting a personal history of CMD had lower adjusted MHLq scores. This finding should be interpreted cautiously because only 23 participants reported a personal history, and the cross-sectional design cannot establish whether mental health experience preceded, followed or altered mental health knowledge. The results suggest that access to understandable interpersonal information may be more relevant to measured MHL than exposure history alone, a hypothesis that warrants prospective investigation.

#### Digital literacy and social media exposure

Digital literacy, rather than any single social-media platform, emerged as the more robust digital correlate of MHL. There was a clear gradient: participants with low, optimal or high digital literacy had lower adjusted MHLq scores than those reporting very high digital literacy. WhatsApp remained one of the most frequently reported information sources, consistent with evidence that university students frequently seek mental health information through social media [41,42], but WhatsApp use itself was not independently associated with the continuous MHLq score after adjustment. Social networking can provide accessible and relatively anonymous channels for information and peer support [43,44]; however, these results favour strengthening digital health literacy and the quality of information across platforms rather than privileging a specific platform. University websites, learning systems and student portals may therefore be useful components of a broader digital MHL strategy.

The construct-validity analyses were broadly consistent with this interpretation. Higher unadjusted scores among students with psychology/psychiatry exposure, later study years, and greater digital literacy were in theoretically expected directions and were mirrored by the adjusted ANCOVA results. Conversely, proximity to mental disorder through a friend or family member did not distinguish MHLq scores in this sample, despite such associations in the original validation and some previous studies [27,31]. The near-zero association with SSQ-14 symptoms suggests that the MHLq is not simply capturing current psychological distress. Taken together, these findings provide partial, rather than comprehensive, evidence of construct validity and reinforce the need to distinguish evidence supporting the overall score from evidence supporting the individual domains.

#### Drug/substance use

Drug/substance use was associated with lower mental health literacy, although this relationship varied by age and should be treated as exploratory given the relatively small subgroup reporting substance use (n = 44). Perceived health status was also associated with the overall MHLq score, but the pattern was non-monotonic: the good-health group scored lower than the very-good-health group, whereas the fair and poor groups did not differ significantly. Consequently, the results do not support a simple interpretation that progressively better perceived health necessarily produces higher MHL. More broadly, prior work has linked health literacy with health-promoting behaviours [45–47]; future studies should examine whether mental health literacy, health behaviours and substance use share common educational or psychosocial pathways.

### Study limitations and strengths

Our study had several limitations. First, convenience sampling within strata may have introduced selection bias and limited generalisability beyond health sciences students at this institution. Second, self-administered measures may be affected by recall and social-desirability bias, and the cross-sectional design precludes causal or temporal inference. Third, the psychometric analyses were secondary and exploratory: the study was not designed as a formal validation study; no independent sample was available for confirmatory factor analysis, and the relatively homogeneous health sciences student population limits generalisability of structural findings. The lower domain reliabilities and incomplete replication of the published factor structure mean that subscale-specific conclusions should be made cautiously. Fourth, the exploratory predictor-selection strategy may produce model instability and should be confirmed in an independent sample; estimates for personal CMD history and drug/substance use are particularly imprecise because these subgroups were small. Fifth, heteroscedasticity was detected, although HC3 robust standard errors and confidence intervals were used to reduce its impact on statistical inference. The age-by-substance-use interaction should also be regarded as hypothesis-generating. Strengths include the large sample size, the use of a standardised MHL measure, an exploratory psychometric evaluation in the local setting, and analysis of the full continuous MHLq total score, which preserves information and avoids imposing an unvalidated high/low threshold.

## Conclusion

The MHLq total score showed good internal consistency and partial support for construct validity among Zimbabwean health sciences students, although domain-level reliability was modest and the published four-domain factor structure was not fully reproduced. These findings support the use of the continuous total MHLq score in the present epidemiological analysis, while cautioning against subscale-specific interpretation and calling for dedicated local validation in future research. We identified variation in mental health literacy by psychology/psychiatry course exposure, digital literacy, year of study, interpersonal information sources, perceived health, substance use and personal CMD history. Campus-wide mental health education and digital-literacy strengthening, including trusted peer/family information pathways, merit prospective evaluation. Longitudinal and intervention studies are needed to establish causal pathways and to determine whether improvements in mental health literacy translate into better help-seeking and mental health outcomes.

## Data Availability

The data are attached as a supplementary file and will be deposited in an online data repository as the paper undergoes peer review.

## Declarations

### Ethics approval and consent to participate

Ethical approval for the study was granted by the University of Zimbabwe Directorate and the Joint Research and Ethics Committee for the University of Zimbabwe, Faculty of Medicine and Health Sciences and Parirenyatwa Group of Hospitals (Ref: JREC/72/2023). All methods were carried out in accordance with the Joint Research and Ethics Committee guidelines and regulations, which conform to the Declaration of Helsinki. Written informed consent was obtained from all participants before enrolment. Unique study identification numbers were used to preserve confidentiality. Data and signed consent forms were stored securely and were accessible only to the research team. Participants could withdraw without penalty.

### Availability of supporting data

The datasets used and analysed during the current study are available and have been uploaded as supporting information files **(S7. Dataset)**.

### CRediT author contribution statement

- **Conceptualization:** Jacqueline Chindondondo, Odette Maungwa, Cathrine Chimutsa, Wendy Makoni, Mazvita Tapfumanei, Alfred Hove, Tariro D. Tunduwani, Sidney Muchemwa, Beatrice K. Shava, Shalom R. Doyce, Anotida R. Hove, Jermaine M. Dambi.
- **Methodology:** Jacqueline Chindondondo, Odette Maungwa, Cathrine Chimutsa, Wendy Makoni, Mazvita Tapfumanei, Alfred Hove, Tariro D. Tunduwani, Sidney Muchemwa, Beatrice K. Shava, Shalom R. Doyce, Anotida R. Hove, Jermaine M. Dambi.
- **Investigation:** Jacqueline Chindondondo, Odette Maungwa, Cathrine Chimutsa, Wendy Makoni, Mazvita Tapfumanei, Alfred Hove, Jermaine M. Dambi.
- **Formal analysis:** Jermaine M. Dambi.
- **Supervision:** Jermaine M. Dambi, Tariro D. Tunduwani, Sidney Muchemwa, Beatrice K. Shava, Shalom R. Doyce, Anotida R. Hove.
- **Project administration:** Jermaine M. Dambi.
- **Writing – original draft:** Jacqueline Chindondondo, Odette Maungwa, Cathrine Chimutsa, Wendy Makoni, Mazvita Tapfumanei, Alfred Hove, Jermaine M. Dambi.
- **Writing – review & editing:** Jacqueline Chindondondo, Odette Maungwa, Tariro D. Tunduwani, Cathrine Chimutsa, Wendy Makoni, Mazvita Tapfumanei, Alfred Hove, Sidney Muchemwa, Beatrice K. Shava, Shalom R. Doyce, Anotida R. Hove, Sandra Mboweni, Webster Mavhu, Dixon Chibanda, Jermaine M. Dambi.
- **Statistical interpretation and critical review:** Jermaine M. Dambi, Sandra Mboweni, Webster Mavhu, Dixon Chibanda.
- All authors reviewed and approved the final manuscript and accept responsibility for their respective contributions.

## Acknowledgments

We thank all participants for their invaluable contributions to this study. Data collection formed part of the undergraduate theses of Jacqueline Chindondondo, Odette Maungwa, Cathrine Chimutsa, Wendy Makoni, Mazvita Tapfumanei and Alfred Hove at the University of Zimbabwe. The undergraduate research was primarily supervised by Jermaine M. Dambi, with co-supervision from Tariro D. Tunduwani, Sidney Muchemwa, Beatrice K. Shava, Shalom R. Doyce and Anotida R. Hove.

## Supporting information captions

**S1 Table. Responses to Mental Health Literacy Questionnaire items (N = 601)**

**S2 Table. Frequencies of responses on the Shona Symptoms Questionnaire (SSQ-14)**.

**S3 Table. Unadjusted associations with the continuous MHLq total score**.

**S4 Table. Internal consistency and factorability of the MHLq in the study sample**.

**S5 Table. Five-factor exploratory factor analysis of MHLq items using principal-axis factoring with oblimin rotation**.

**S6 Table. Hypothesis-based construct validity of the MHLq total score**.

**S7. Dataset**

